# Seroprevalence of antibodies against SARS-CoV-2 among health care workers in a large Spanish reference hospital

**DOI:** 10.1101/2020.04.27.20082289

**Authors:** Alberto L Garcia-Basteiro, Gemma Moncunill, Marta Tortajada, Marta Vidal, Caterina Guinovart, Alfons Jiménez, Rebeca Santano, Sergi Sanz, Susana Méndez, Anna Llupià, Ruth Aguilar, Selena Alonso, Diana Barrios, Carlo Carolis, Pau Cisteró, Eugenia Chóliz, Angeline Cruz, Silvia Folchs, Chenjerai Jairoce, Jochen Hecht, Montserrat Lamoglia, Mikel J. Martínez, Robert A. Mitchell, Natalia Ortega, Nuria Pey, Laura Puyol, Marta Ribes, Neus Rosell, Patricia Sotomayor, Sara Torres, Sarah Williams, Sonia Barroso, Anna Vilella, José Muñoz, Antoni Trilla, Pilar Varela, Alfredo Mayor, Carlota Dobaño

## Abstract

**Background:** Health care workers (HCW) are a high-risk population to acquire SARS-CoV-2 infection from patients or other fellow HCW. At the same time, they can be contagious to highly vulnerable individuals seeking health care. This study aims at estimating the seroprevalence of antibodies against SARS-CoV-2 and associated factors in HCW from a large referral hospital in Barcelona, Spain, one of the countries hardest hit by COVID-19 in the world.

**Methods:** From 28 March to 9 April 2020, we recruited a random sample of 578 HCW from the human resources database of Hospital Clínic in Barcelona. We collected a nasopharyngeal swab for direct SARS-CoV-2 detection through real time reverse-transcriptase polymerase chain reaction (rRT-PCR), as well as blood for plasma antibody quantification. IgM, IgG and IgA antibodies to the receptor-binding domain of the spike protein were measured by Luminex. The cumulative prevalence of infection (past or current) was defined by a positive SARS-CoV-2 rRT-PCR and/or antibody seropositivity.

**Results:** Of the 578 total participants, 39 (6.7%, 95% CI: 4.8-9.1) had been previously diagnosed with COVID-19 by rRT-PCR, 14 (2.4%, 95% CI: 1.4-4.3) had a positive rRT-PCR at recruitment, and 54 (9.3%, 95% CI: 7.2-12.0) were seropositive for IgM and/or IgG and/or IgA against SARS-CoV-2. Of the 54 seropositive HCW, 21 (38.9%) had not been previously diagnosed with COVID-19, although 10 of them (47.6%) reported past COVID-19-compatible symptoms. The cumulative prevalence of SARS-CoV-2 infection was 11.2% (65/578, 95% CI: 8.9-14.1). Among those with evidence of past or current infection, 40.0% (26/65) had not been previously diagnosed with COVID-19, of which 46.2% (12/26) had history of COVID-19-compatible symptoms. The odds of being seropositive was higher in participants who reported any COVID-19 symptom (OR: 8.84, 95% CI: 4.41-17.73). IgM levels positively correlated with age (rho=0.36, p-value=0.031) and were higher in participants with more than 10 days since onset of symptoms (p-value=0.022), and IgA levels were higher in symptomatic than asymptomatic subjects (p-value=0.041).

**Conclusions:** The seroprevalence of antibodies against SARS-CoV-2 among HCW was lower than expected. Thus, being a high-risk population, we anticipate these estimates to be an upper limit to the seroprevalence of the general population. Forty per cent of those with past or present infection had not been previously diagnosed with COVID-19, which calls for active periodic rRT-PCR testing among all HCW to minimize potential risk of hospital-acquired SARS-CoV-2 infections.

## Introduction

COVID-19 is a novel viral disease caused by SARS-CoV-2 that was first detected in Wuhan, China, in December 2019.^1^ Given the alarming levels of spread, severity of disease, and number of affected countries, the World Health Organization (WHO) declared COVID-19 as a pandemic on March 11^th^, 2020.^2^ The clinical syndrome caused by SARS-CoV-2 ranges from very mild symptomatology to severe pneumonia, acute respiratory distress syndrome (ARDS) and death.^3^ However, several reports show that many individuals might carry the virus without presenting any symptoms for several weeks.^4–6^ Thus, the exact number of individuals who have been infected by SARS-CoV-2 is currently unknown.

Health care workers (HCW) are the frontline workforce for clinical care of suspected and confirmed COVID-19 cases. Consequently, they are presumably exposed to a higher risk of acquiring the disease than the general population and, if infected, pose a risk to vulnerable patients and fellow HCW.^7^ In a tertiary hospital in Madrid, Spain (one of the regions with the highest COVID-19 attack rates in the country), 38% (791/2085) of HCW tested positive for SARS-CoV-2 by real time reverse-transcriptase polymerase chain reaction (rRT-PCR) in March 2020 (11.6% of that hospital population).^8^ HCW with a positive rRT-PCR diagnosis need to be isolated, and their close contacts –many of them co-workers– should be quarantined. Thus, if transmission rises, the number of frontline HCW could become insufficient to respond to the healthcare demand. To cope with this scenario, several strategies, including periodic screenings, weekly-shifts and other organizational measures are being implemented in a variety of settings to guarantee proper patient care.^9^ Nonetheless, quantification and characterization of SARS-CoV-2 infection within health care facilities is unknown in most countries hard-hit by the COVID-19 epidemic.

Seroprevalence studies can provide relevant information on the proportion of people who have experienced a recent or past infection. They are relevant when conducted in the community, but also for critical population subgroups such as nursing homes or health care facilities. Monitoring the prevalence of infection among HCW (regardless of history of symptoms) is useful for assessing the level of exposure among hospital personnel and identifying high risk departments. Likewise, knowledge of past infection among HCW could be useful for avoiding unnecessary quarantines and for health care resource planning.^10^ Although there is a growing body of evidence on the immunological responses against SARS-CoV-2, the time to seroconversion and the antibody levels elicited are not well characterized yet. Importantly, the correlation between seropositivity or antibody levels and protection against reinfection, as well as the duration of protective immunity, remains to be elucidated.^11^

This study aims to estimate the seroprevalence of antibodies against SARS-CoV-2 and characterize the antibody profile in HCW from Hospital Clínic of Barcelona, one of the reference centers in Spain for the diagnosis and treatment of COVID-19 disease. As a secondary objective, we aim to assess the overall infection prevalence (past and current) to SARS-CoV-2 as well as the prevalence of asymptomatic infections.

## Methods

### Study design, population and setting

The study design consists of a series of four cross-sectional surveys (at baseline, 1 month, 6 months and 12 months) in a cohort of randomly selected HCW from Hospital Clínic of Barcelona (HCB). We hereby present the first cross-sectional survey, conducted from March 28^th^ to April 9^th^, 2020. The study population was defined as those who deliver care and services to patients, either directly as physicians or nurses, or indirectly as assistants, technicians, stretcher-bearers or other support staff (administrative officers, cleaning, kitchen, laundry, maintenance, etc.).^12^ Inclusion criteria included being an adult (>17 years) worker at HCB registered at the Human Resources department. Exclusion criteria included: a) absenteeism from workplace in the last 30 days (i.e. on vacation, sick leave, sabbatical), b) working exclusively outside the HCB or Maternity main buildings with no interaction with patients on a daily basis, c) retirement or end-of-contract planned within one year after the recruitment date, and d) participating in COVID-19 clinical trials for preventive or treatment therapies.

HCB is a large University of Barcelona teaching hospital. With over 700 beds, it is the main public supplier of specialized health services for a population of around 540,000 inhabitants and also acts as a tertiary referral hospital.^13^

### Procedures

A random sample of HCW was selected from the HCB’s Human Resources database (as of March 9^th^, 2020). Selected individuals were approached telephonically following the order of the random list were excluded upon review of inclusion and exclusion criteria or after 3 phone calls (different days) without response.

After obtaining written informed consent, we filled out a standardized electronic questionnaire programmed in REDCap (Research Electronic Data Capture)^14^ for each participant, with the following information: demographics (age, sex, household size, etc.), professional information (occupation, hospital department, shift), clinical information such as history of COVID-19-compatible symptoms during the previous month (cough, sore throat, runny nose, fatigue, shortness of breath, fever, headache, vomiting, diarrhea, anosmia, ageusia and chills) date of onset and resolution of symptoms, history of rRT-PCR testing, comorbidities, and history of close contact with COVID-19 cases.

We collected a nasopharyngeal swab (DeltaLabs ref: 304273) for the detection of SARS-CoV-2 RNA by rRT-PCR and a venous blood draw for immunological assessments. Both procedures were performed by trained nurses using appropriate personal protective equipment (PPE). Samples were transported to the laboratory within 3 hours of sample collection. Nasopharyngeal swabs and plasma samples were stored at −80°C until analysis.

For participants reporting to be isolated at home (i.e. due to a COVID-19 diagnosis) or on quarantine, data and specimen collection took place at their households following the relevant biosafety protocols.

### Laboratory procedures

#### rRT-PCR

After adding 500 μl of Zymo DNA/RNA Shield Lysis Buffer to the same amount of nasopharyngeal sample collection media, RNA was extracted using the Quick-DNA/RNA Viral MagBead kit (Zymo) and the TECAN Dreamprep robot. Five microliters of RNA solution were added to 15 μl of rRT-PCR master mix (Luna Universal Probe One-Step RT-qPCR Kit; New England Biolabs) and used for amplification of SARS-CoV-2 N1 and N2 regions, as well as the human RNase P gene as control, using probes, primers and cycling conditions described in the CDC-006–00019 CDC/DDID/NCIRD/ Division of Viral Diseases protocol (3/30/2020 release). Each batch of RNA extractions and rRT-PCR reactions included 3 positive controls (EURM-019 single stranded RNA fragments of SARS-CoV-2 provided by the European Commission Joint Research Centre), 2019-nCoV_N_Positive Control (IDT integrated technologies, ref. 10006625) and Hs_RPP30 Positive Control (IDT integrated technologies, ref. 10006626), as well as negative controls. A positive result was considered if the Ct values for N1, N2 and RNase P were below 40. Samples discordant for N1 and N2 were repeated and samples with a Ct≥40 for RNase P were considered as invalid.

#### Quantification of antibodies to SARS-CoV-2 by Luminex

To establish seroprevalence, we used a serological assay based on the Luminex technique that has the benefit of a higher dynamic range than other assays, favoring the quantification of immunoglobulin levels. We measured antibodies against the Receptor-Binding Domain (RBD) of the spike glycoprotein of SARS-CoV-2, which is, together with the nucleocapsid protein (NP), one of the most immunogenic antigens.^15^ Antibodies to RBD correlate with neutralizing antibodies^16,17^ that could be associated with protection based on studies of other coronaviruses and animal models.^17–20^ The RBD antigen, kindly donated by the Krammer lab (Mount Sinai, New York)^21^, was coupled to magnetic MAGPLEX 6.5 μm COOH-microspheres from Luminex Corporation (Austin, TX) at a concentration of 40 μg/ml for 10,000 beads/μl, as previously described^22^.

Antigen-coupled beads were added to a 96-well μClear® flat bottom plate (Greiner Bio-One, 655096) at 2000 beads/well in a volume of 90 μL/well of phosphate buffered saline + 1% bovine serum albumin + 0.05% sodium azide (PBS-BN). Next, 10 μl of test plasma samples (final dilution 1/500), 10 μl of a positive control (pool of 20 plasmas from subjects with a positive SARS-CoV-2 rRT-PCR, at four dilutions, 1/500, 1/2000, 1/8000 and 1/32000, for QA/QC), and 10 μl of 2 negative controls (plasmas from European subjects collected before the COVID-19 pandemic, at 1/500), were added per plate. Two blank control wells with beads in PBS-BN were set up to measure background signal. Plates were incubated at room temperature (RT) for 2 h on a microplate shaker at 500 rpm and protected from light. Plates were washed three times with 300 μl/well of PBS-Tween20 0.05%, using a magnetic manual washer (Millipore, 43–285). A hundred microliters of biotinylated secondary antibody diluted in PBS-BN (anti-human IgG, B1140, 1/1250; anti-human IgM, B1265, 1/1000; or anti-human IgA, SAB3701227, 1/500; Sigma) were added to all wells and incubated for 45 min at 500 rpm at RT and protected from light. Plates were washed three times and 100 μL of streptavidin-R-phycoerythrin (Sigma, 42250) diluted 1:1000 in PBS-BN were added and incubated during 30 min at 500 rpm, RT and protected from light. Plates were washed three times, and beads resuspended in 100 μl of PBSBN and kept overnight at 4°C, protected from light. The next day, plates were read using a Luminex xMAP® 100/200 analyzer with 70 μl of acquisition volume per well, DD gate 5000-25000 settings, and high PMT option. At least 50 beads were acquired per sample. Crude median fluorescent intensities (MFI) were exported using the xPONENT software. Assay cutoff was calculated as 10 to the mean plus 3 standard deviations of log_10_-transformed MFIs of 47 negative controls. Sensitivity of the assay using samples from participants previously diagnosed with COVID-19 and with more than 10 days since the onset of symptoms was 97% for IgA and IgG and 75% for IgM, with specificities of 100% for IgG and IgM and 98% for IgA (**Table S1**). The area under the receiver operating characteristic curve (AUC) was >0.97 for each of the isotypes using these same samples (**Figure S1A**) and >0.87 using samples from any participant previously diagnosed with COVID-19 regardless of the time since onset of symptoms (**Figure S1B**).

#### Sample size and statistical analysis

In order to assess the seroprevalence against SARS-CoV-2 at two time points (month 0 and month 1), with a precision of 5% and a 95% confidence interval (CI), a loss to follow up between month 0 and month 1 of 5% and assuming that the prevalence at month 0 was 30% and at month 1 was 50%, with a finite population, we estimated we would need 570 HCW. Given the uncertainty about what the seroprevalence would be at month 1, we used 50%, which provides the most conservative sample size.

Seroprevalence of antibodies against SARS-CoV-2, prevalence of SARS-CoV-2 infection by rRT-PCR, and cumulative prevalence of past or current infection (positive SARS-CoV-2 rRT-PCR and/or antibody seropositivity), were calculated as proportions with 95% CI. We have tested the association between variables with Chi-square or Fisher’s exact test (for categorical variables) test and T-Student test (for continuous quantitative variables). Univariable and multivariable logistic regression models (MLM) were run to evaluate factors associated with seroprevalence of antibodies against SARS-CoV-2. For the variables to be included in the MLM model, we used a stepwise selection, starting with the full model, and using a p-value of 0.10 for removal and 0.05 for addition of variables. A diagnosis of COVID-19 was excluded from the MLM because it was assumed to be the source for antibody generation and the high expected correlation with COVID-19 symptoms reported.

Spearman correlations were performed to assess the association of antibody levels with age. Wilcoxon Sum Rank test was used to compare the antibody levels between different groups. Receiver Operating Characteristic (ROC) curves and their correspondent AUC were calculated using the predicted values estimated by logistic regression models with MFI for IgM, IgG, IgA or their combination as predictors and the rRT-PCR result as outcome. The analysis was carried out using the statistical software Stata v16.1 (College Station, TX: StataCorp LLC) and R studio version R-3.5.1 (packages used: ggplot2 and pROC).

### Ethical considerations

The protocol and informed consent form were reviewed and approved by the Institutional Review Board (IRB) at HCB (CEIm) prior to study implementation (Ref number: HCB/2020/0336).

### Data availability

Anonymized data will be available upon request to corresponding author and approval by HCB’s Ethics Committee (CEIm). If approved, no restriction to data access.

### Code availability

We will provide the code for logistic regression and figures in supplementary material. Other codes are available upon request to the corresponding author.

## Results

### Baseline characteristics

From a total number of 5598 HCW registered at HCB as of March 9^th^ 2020, we approached the first 1172 randomly selected individuals, following the order of the list. Of these, 798 were eligible to participate and 583 were recruited, yielding a participation rate of 74.3%. We then excluded 5 recruited participants after re-checking inclusion and exclusion criteria (**Figure 1**). A total of 578 participants were included in the analysis, of whom 314 (54.3%) were younger than <45 years of age.

**Figure 1:**
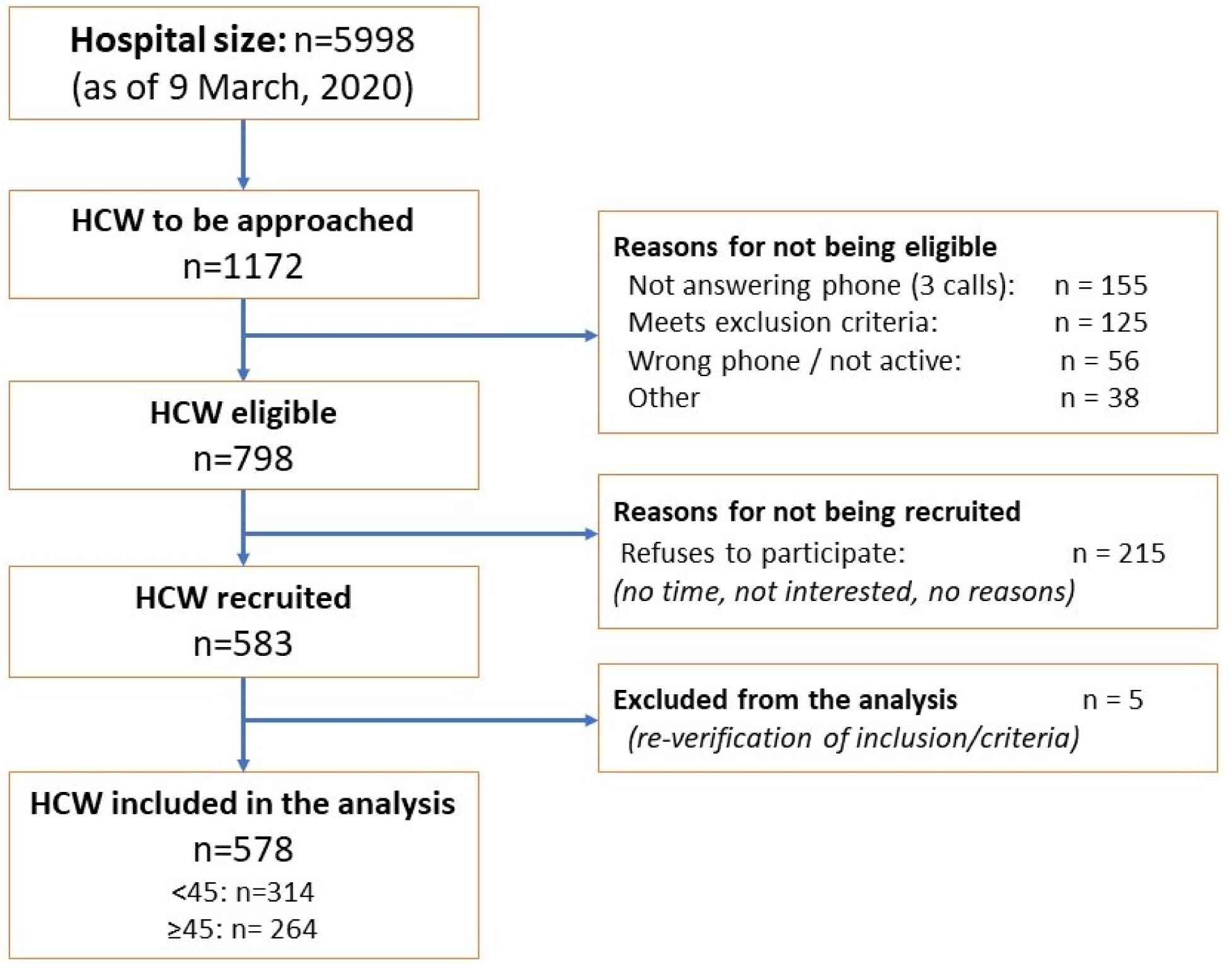
Study participant flowchart.

The mean age of participants was 42.1 years (SD: 11.6) and 72.1% were female. Around half (288/578, 49.8%) were nurses, auxiliary nurses and stretcher-bearers and 25.4% (147/578) were physicians. Eleven per cent of the participants reported having comorbidities and 36.3% reported having had COVID-19-compatible symptoms in the previous month. Thirty-nine participants (6.7%) had been previously diagnosed with COVID-19 confirmed by rRT-PCR, of which only one had required hospital admission (**Table 1**).

**Table 1.** Baseline characteristics of study participants.

|  |  | **<45y**<br>**(n=314)** | **>=45y**<br>**(n=264)** | **Total** | **P-value** |
| --- | --- | --- | --- | --- | --- |
| **Sex**<sup>1</sup> | *Male* | 88 (28%) | 73 (28%) | 161 (28%) | 0.9204<sup>2</sup> |
|  | *Female* | 226 (72%) | 191 (72%) | 417 (72%) |  |
| **Professional category**<sup>1</sup> | *Nurse / Auxiliary nurse /*<br>*Stretcher-bearer* | 166 (53%) | 122 (46%) | 288 (50%) | 0.0020<sup>2</sup> |
|  | *Physician* | 86 (27%) | 61 (23%) | 147 (25%) |  |
|  | *Lab technologist / other tech.* | 26 (8%) | 19 (7%) | 45 (8%) |  |
|  | *Administrative officers, Other*<sup>6</sup> | 36 (11%) | 62 (23%) | 98 (17%) |  |
| **Age**<sup>3</sup> |  | 33 (6.6) | 52,9 (5.3) | 42,1 (11.6) |  |
| **Daily contact with patients** | *No* | 55 (18%) | 68 (26%) | 123 (21%) | 0.0159<sup>2</sup> |
|  | *Yes* | 259(82%) | 196 (74%) | 455 (79%) |  |
| **Working in a COVID19 unit**<sup>1</sup> | *No* | 151 (48%) | 164 (62%) | 315 (54%) | 0.0007<sup>2</sup> |
|  | *Yes* | 163 (52%) | 100 (38%) | 263 (46%) |  |
| **Close contact with COVID19**<br>**confirmed or suspected case**<sup>1</sup> | *No* | 65 (21%) | 72 (27%) | 137 (24%) | 0.0642<sup>2</sup> |
|  | *Yes* | 249 (79%) | 192 (73%) | 441 (76%) |  |
| **Previously diagnosed with COVID19**<br>**by rRT-PCR**<sup>1</sup> | *No* | 292 (93%) | 247 (94%) | 539 (93%) | 0.7866<sup>2</sup> |
|  | *Yes* | 22 (7%) | 17 (6%) | 39 (7%) |  |
| **Comorbidities**<sup>1*</sup> | *No* | 292 (93%) | 225 (85%) | 517 (89%) | 0.0025<sup>2</sup> |
|  | *Yes* | 22 (7%) | 39 (15%) | 61 (11%) |  |
| **Household size**<sup>3</sup> |  | 2.6 (1.1) | 2.9 (1.2) | 2.8 (1.2) | 0.0200<sup>4</sup> |
| **Received Flu vaccine (2019-2020**<br>**season)**<sup>1</sup> | *No* | 175 (56%) | 164 (62%) | 339 (59%) | 0.1203<sup>2</sup> |
|  | *Yes* | 139 (44%) | 100 (38%) | 239 (41%) |  |
| **Reporting COVID-19 compatible**<br>**symptoms within previous month**<sup>1</sup> | *No* | 196 (62%) | 172 (65%) | 368 (64%) | 0.4965<sup>2</sup> |
|  | *Yes* | 118 (38%) | 92 (35%) | 210 (36%) |  |

1: n (Column percentage)
2: Chi-squared test
3: Arithmetic Mean (SD) [n]
4: t-test
5: Fisher's exact test
6. Includes, cleaning, kitchen and maintenance staff
\* Comorbidities include: heart and liver disease, diabetes, chronic respiratory and renal disease, cancers and autoimmune and other immunological disorders.

### Prevalence of current infection as determined by a positive rRT-PCR

Fourteen participants (2.6%, 95% CI: 1.4-4.3%) had a positive rRT-PCR of SARS-CoV-2 at the time of recruitment and 43 (7.4%) had an invalid rRT-PCR result. Among participants with a positive rRT-PCR at recruitment, 8 of 14 (57.1%) had a previous COVID-19 diagnosis, and 3 of 14 (21.4%) did not report any COVID-19-compatible symptom in the previous month. Only 1 of the 6 participants with positive rRT-PCR at recruitment and no history of previous COVID-19 diagnosis had detectable antibodies.

### Seroprevalence of antibodies against SARS-CoV-2

Fifty-four participants (9.3%, 95% CI: 7.2-12.0) were seropositive for IgM and/or IgG and/or IgA against SARS-CoV-2 (**Table 2**). A total of 36 (6.2%), 44 (7.6%) and 47 (8.1%) participants were positive for IgM, IgG and IgA, respectively (**Figure 2**). Four participants were seropositive for IgM only (IgG- & IgA-), 2 were seropositive for IgG only (IgM- & IgA-) and 5 were seropositive for IgA only (IgG- & IgM-). Around 15% (6/39) of HCW who had been previously diagnosed with COVID-19 by rRT-PCR did not show a detectable response of any of the antibody isotypes (**Table S2**). However, the days since onset of symptoms to recruitment were less than 10 in 4 out of the 6 individuals without detectable antibodies and one individual had no symptoms. Twenty per cent (11/54) of seropositive participants did not report COVID-19 compatible symptoms in the previous month. Over 39% (21/54) of seropositive HCW had never been diagnosed of COVID-19, although 10 of these (47.6%) reported past COVID-19-compatible symptoms.

**Table 2.** Overall proportion of HCW with a) detectable antibodies b) history of past positive rRT-PCR, c) Positive rRT-PCR at study recruitment, and d) Cumulative prevalence of infection (past/current rRT-PCR and/or antibodies).

|  | *n* | *Total* | *% (95% CI)* | *Not previously*<br>*diagnosed as*<br>*COVID-19 by*<br>*rRT-PCR (n (%))* | *COVID19-*<br>*symptoms*<br>*reported\* n (%)* |
| --- | --- | --- | --- | --- | --- |
| Seropositive to SARS Cov-2<br>Antibodies (IgA and/or IgM and/or<br>IgG) | 54 | 578 | 9.3% (7.1-12.0) | 21 (38.9%) | 10 (47.6%) |
| History of past positive rRT-PCR | 39 | 578 | 6.7% (4.8-9.1) | 0 (0%) | 0 (0%) |
| Positive rRT-PCR at study<br>recruitment** | 14 | 535 | 2.6% (1.4-4.3) | 6 (42.9%) | 3 (50.0%) |
| Any evidence of past/current<br>infection by rRT-PCR or serology | 65 | 578 | 11.2% (8.8-14.1) | 26 (40.0%) | 12 (46.2%) |

\*Among those not previously diagnosed as COVID-19 (from previous column)
\*\*Results of 43 of the 578 rRT-PCRs done were invalid (Ct≥40 for RNase P)

**Figure 2:**
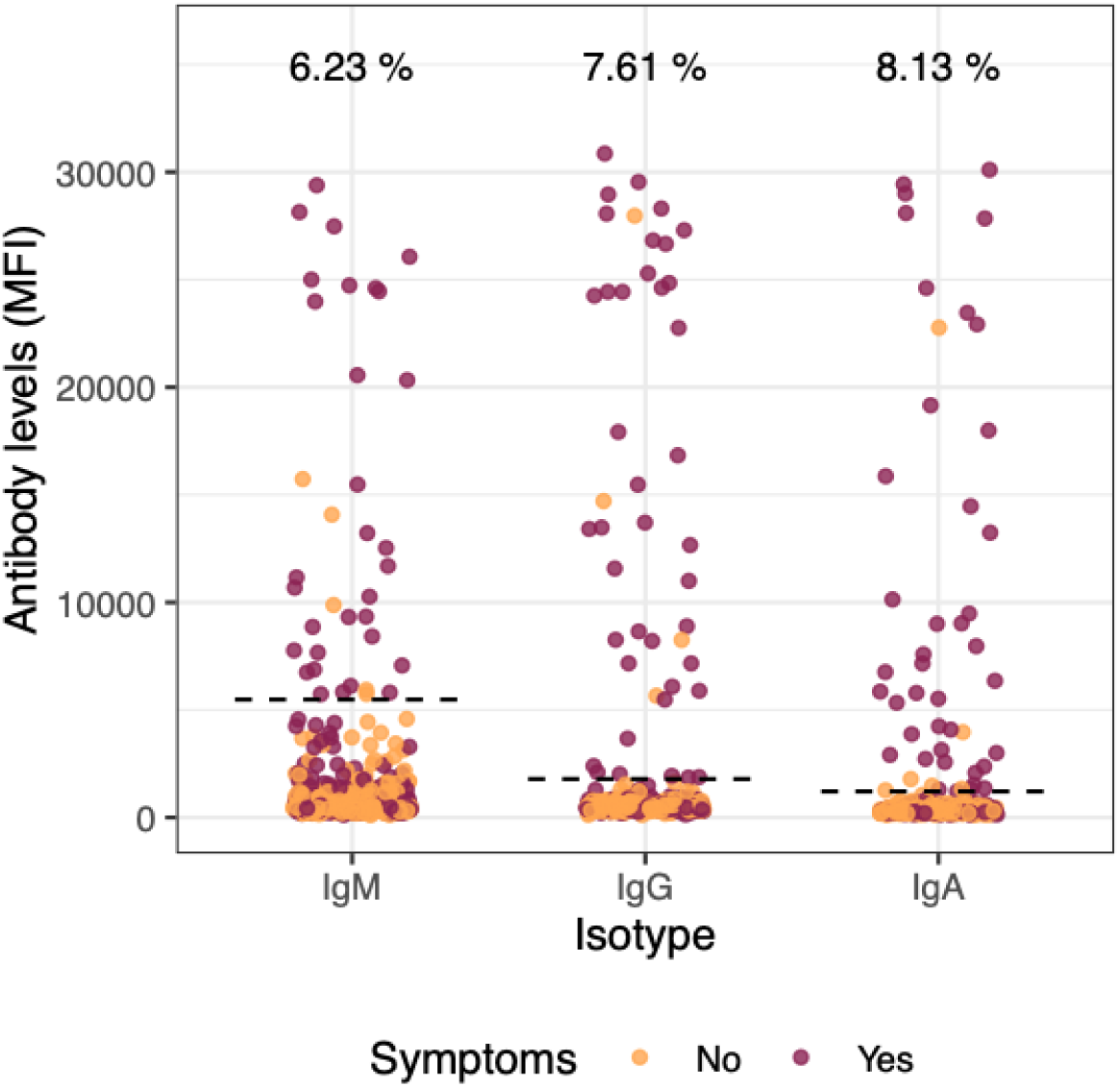
SARS-CoV-2 antibody levels in all study participants. Dots depict the levels (median fluorescence intensity, MFI) of IgM, IgG, and IgA against Receptor Binding Domain of the SARS-CoV-2 Spike glycoprotein. Dashed lines indicate the seropositivity threshold calculated with pre-pandemic controls as the 10 to the mean plus 3 standard deviations of log_10_-transformed MFIs). The percentage of seropositive subjects is shown for each antibody isotype. Orange and burgundy dots show subjects who did not have or did have history of at least one COVID-19 compatible symptom, respectively. N=578.

The odds of being seropositive were higher in participants who reported having had any COVID-19-compatible symptom in the previous months (adjusted OR: 8.84, 95% CI 4.41-17.73, p<0.0001) (**Table 3**). The individual symptoms more strongly associated with seropositivity were (in order): anosmia (OR: 83.0, 95%CI: 29.6-232.9), ageusia (OR: 71.4, 95% CI: 25.4-200.8), fever (OR: 11.4, 95% CI: 6.0-31.3) and fatigue (OR: 11.2, 95% CI: 6.1-20.7), all of them with a p<0.0001 in the univariable analysis. There was some evidence in the MLM that those with higher household size had higher seroprevalence (OR for every additional household member: 1.25; 95% CI: 0.96-1.62; p=0.09). The professional category, working in COVID-19 units, daily contact with patients, close contact with a COVID-19 case, comorbidities or sex, did not show any statistically significant association with presence of antibodies to SARS-CoV-2 (**Table 3, Figure S2**).

**Table 3.**
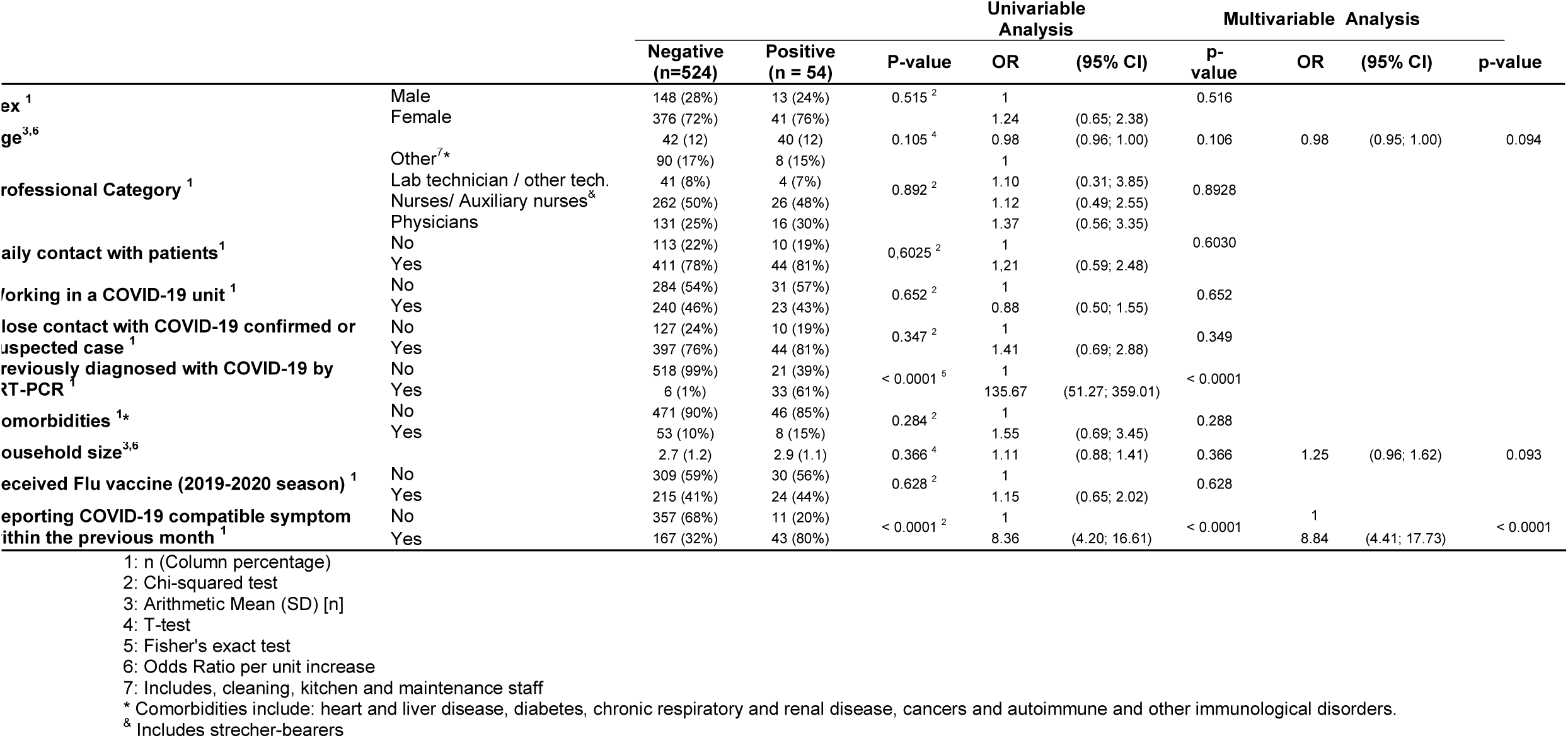
Univariable and multivariable analysis of factors associated with having detectable antibodies (IgM and/or IgG and/or IgA)

Among seropositive HCW, there were no statistically significant associations of antibody levels with sex (**Figure 3A**). IgM levels positively correlated with age (rho=0.36, p=0.031) (**Figure 3B**). IgA levels were higher in participants reporting COVID-19-compatible symptoms in the previous month than in those reporting being asymptomatic (p=0.041) (**Figure 3C**), and among symptomatic individuals, duration of symptoms greater than 10 days was associated with higher IgM levels (p=0.021) (**Figure 3D**).

**Figure 3:**
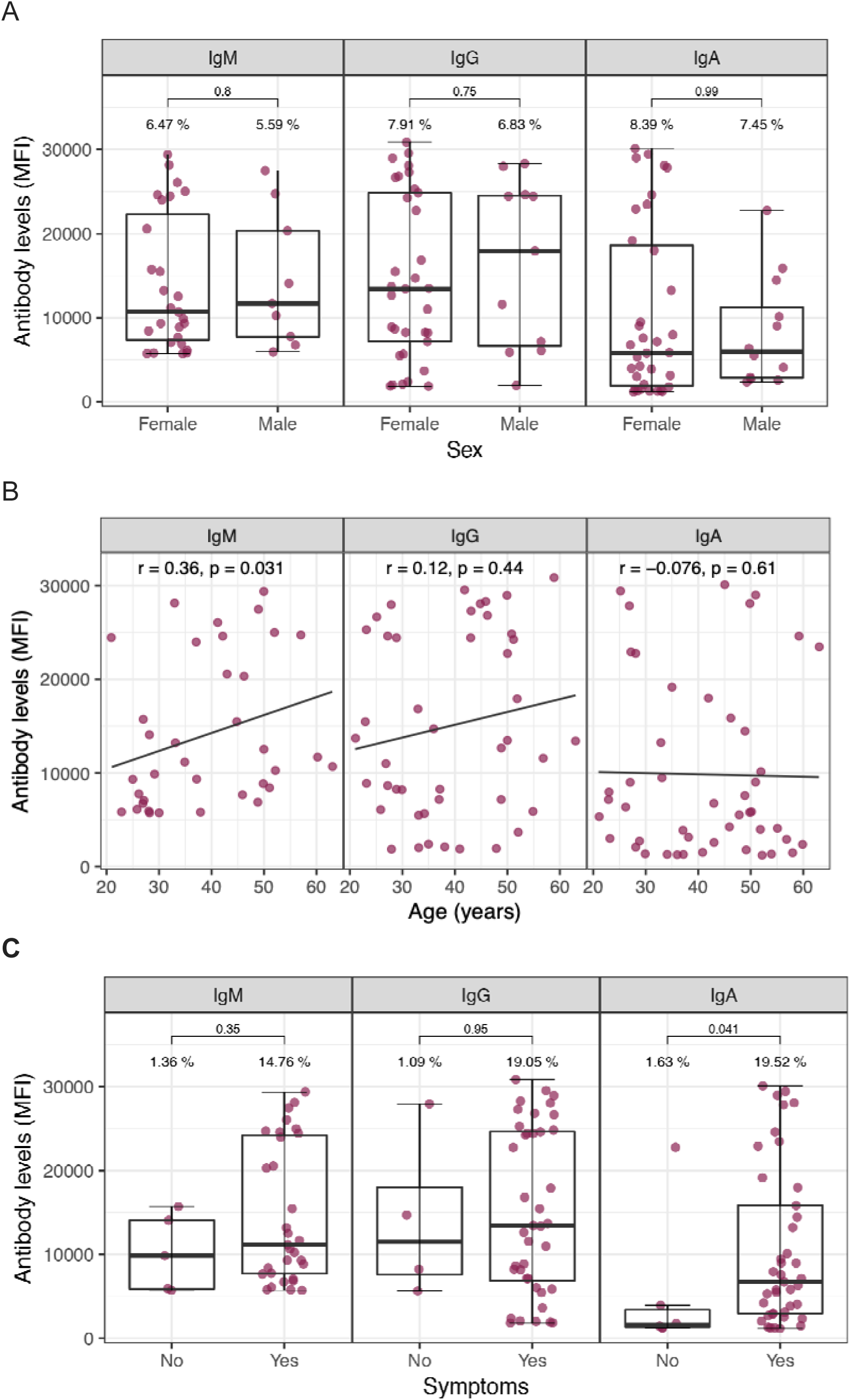
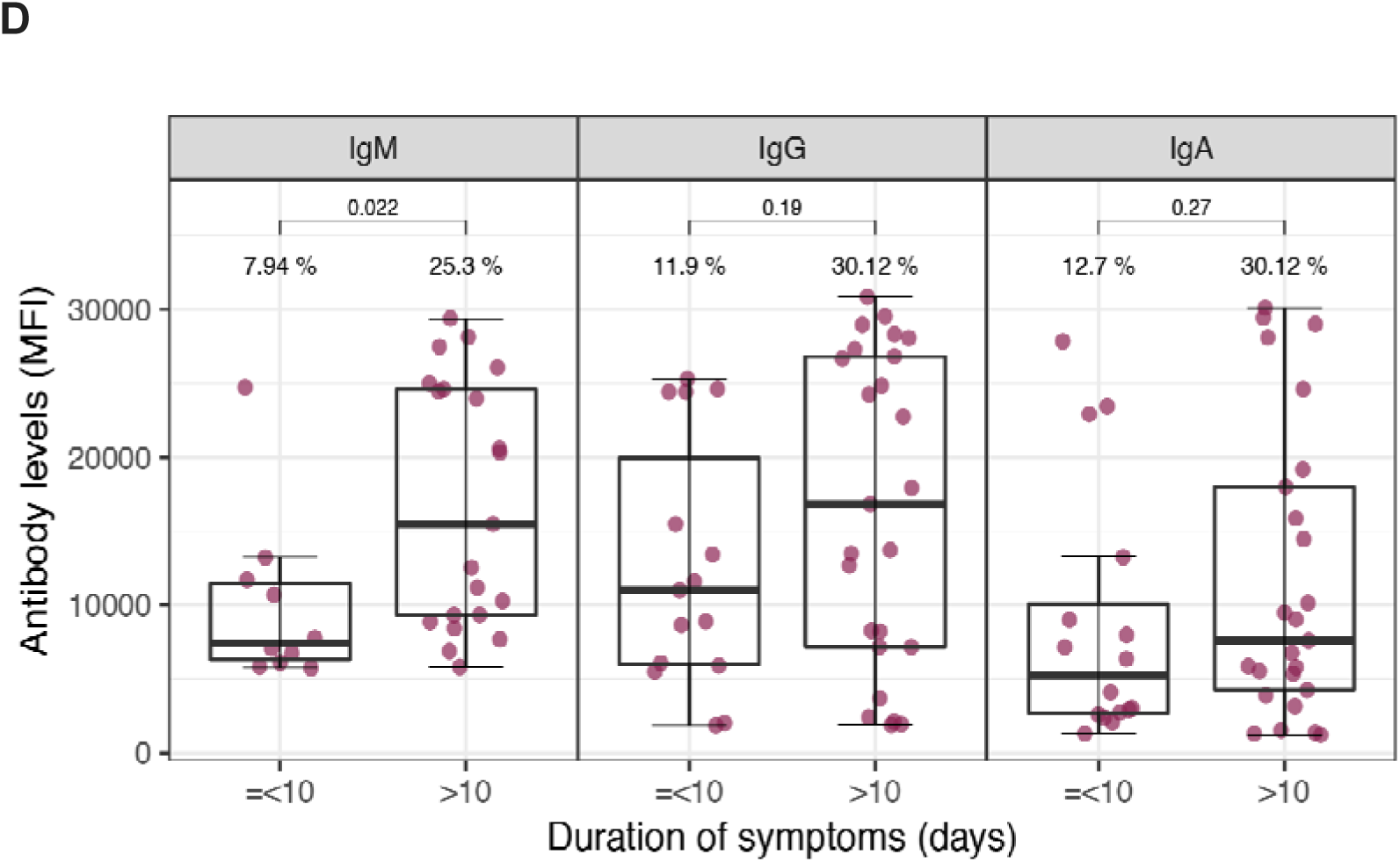
SARS-CoV-2 antibody levels by demographic and clinical variables. Levels (median fluorescence intensity, MFI) of IgM, IgG and IgA against Receptor Binding Domain of the SARS-CoV-2 Spike glycoprotein by sex (A), age (B), symptoms (C), and duration of symptoms (D). For A-C, data are shown only for seropositive subjects for IgM (N=36), for IgG (N=44), and for IgA (N=47). For D, data are shown only for seropositive and symptomatic subjects for IgM (N= 31), for IgG (N=40), and for IgA (N=41). Percentages indicate the proportion of seropositive subjects within each category of the x axis. Boxes depict median MFIs and interquartile ranges (IQR); the lower and upper hinges correspond to the first and third quartiles; whiskers extend from the hinge to the highest or lowest value within 1.5 × IQR of the respective hinge. Wilcoxon rank test was used to assess statistically significant differences in antibody levels between groups in (A), (C) and (D). Spearman test was used to assess the correlations in (B).

Among HCW reporting symptoms in the last month, antibodies were detected in individuals with 6 or more days between symptoms onset and recruitment for IgA and later for IgM and IgG (**Figure 4**), with no seropositive results detected among individuals with symptoms onset less than 6 days prior to the recruitment visit. In fact, we only detected antibodies in two participants surveyed less than 10 days after onset of symptoms (**Table S3**). Antibody levels increased and peaked between day 20 and 25 for IgM and IgG, and a few days earlier for IgA. There were only three seropositive HCW with symptoms onset having occurred earlier than 25 days prior to survey (**Table S3**).

**Figure 4:**
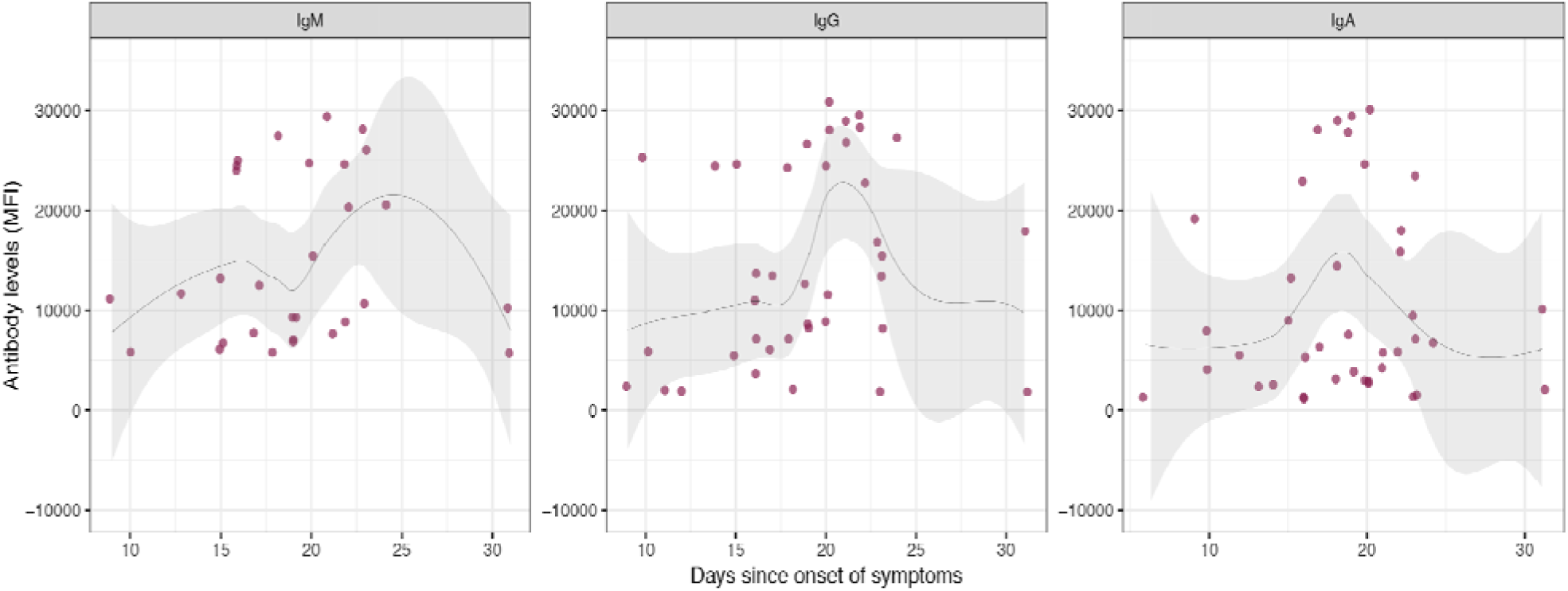
SARS-CoV-2 antibody levels by time since onset of symptoms in seropositive subjects. Levels (median fluorescence intensity, MFI) of IgM (A), IgG (B) and IgA (C) against Receptor Binding Domain of the SARS-CoV-2 Spike glycoprotein by days since onset of any symptom. Data are shown only for seropositive subjects with any symptom compatible with COVID-19 (n=30 for IgM, 39 for IgG, and 40 for IgA). The fitting curve was calculated using the LOESS (locally estimated scatterplot smoothing) method. Shaded areas represent 95% CIs. One subject seropositive for the three isotypes and who started symptoms 40 days before serological testing was not shown.

### Overall cumulative prevalence of past or current infection

Sixty-five HCW had either a positive rRT-PCR in the past or at survey recruitment, or had a positive antibody response (**Table 2**). Thus, the cumulative prevalence of SARS-CoV-2 infection was 11.2% (95% CI: 8.9-14.1). Among them, 23.1% (15/65) did not report any COVID-compatible symptom in the previous month. Forty per cent (26/65) had not been previously diagnosed with COVID-19, although 12 of them reported COVID-19 compatible symptoms.

## Discussion

This is, to our knowledge, the first study reporting seroprevalence of antibodies against SARS-CoV-2 among a representative sample of HCW in a COVID-19 high burden country. We found that 9.3% (95% CI: 7.2-12.0) of HCW from a large Spanish referral hospital (recruited from March 28^th^ to April 9^th^, 2020) developed detectable IgA, IgG and/or IgM antibodies. Given that HCW are a high risk population for SARS-CoV-2, it is likely that the community seroprevalence is lower than this figure, showing that we are still very far from reaching the 67% herd immunity level that is estimated to be needed to protect the susceptible population,^23^ assuming that this immunity prevents from reinfection. The seroprevalence found was lower than expected, based on the large number of rRT-PCR positive cases reported in a referral hospital in Madrid in March 2020 (11.6% of all hospital workers),^8^ and an estimate from modeling studies of 15% seroprevalence for the overall Spanish population in March 2020.^24^ The likely higher availability of PPE compared to other hospitals, and the early implementation of rRT-PCR screening programs in HCW working in COVID-19 units, coupled with timely case identification and effective contact tracing and quarantines for those outside COVID-19 unit, could explain a relatively low number of infections in our study.

Combining data from antibody detection and previous or current positive rRT-PCR, the cumulative prevalence of SARS-CoV-2 infection rose to 11.2%. However, 40.0% of the seropositive HCW had not previously been diagnosed with COVID-19 and 23.1% were asymptomatic, indicating that a large percentage of infections with mild or no symptoms, as previously described.^25^ This calls for early detection/screening programs to be broadly and timely implemented in HCW to decrease in-hospital transmission as well as reinforce the critical role of PPE usage.^26^

Seropositivity was higher in participants who reported having had any COVID-19 symptom within the last month (OR: 8.84) and 80% of seropositive HCW did report having had symptoms. Although most COVID-19 symptoms are common to most other upper respiratory viral infections, those more highly associated with seropositivity were by far anosmia and ageusia (both OR>70) that, although infrequent, seem to be quite specific for COVID-19.^27,28^ As expected, having developed the disease was the most important factor associated with the development of antibodies (OR: 135.6). In addition, there was some evidence that the higher the size of the household, the higher the odds (OR: 1.25) of being seropositive (p<0.09), potentially because household exposure is an added source of infection among HCW. None of the professional categories or being directly involved in clinical care were factors associated with higher odds of being seropositive. Working in a COVID-19 unit was also not associated with seropositivity, which might be explained by a higher perception of risk leading to a better protection with PPEs, more careful practices and thus, a lower risk of acquiring the infection.^29^ Nonetheless, the relatively low number of seropositive HCW in our sample hinders any firm conclusion about associations between professional categories, level of patient interaction, and risk of infection.

Using only the RBD antigen in the assay but three different isotypes, we could detect antibodies in 97% of participants with a previous positive rRT-PCR and more than 10 days since onset of symptoms. This is in line with previous reports showing that seroconversion occurs between 2-3 weeks after onset of symptoms.^11^ Importantly, we detected lower IgA levels in seropositive participants without symptoms, in line with a previous observation of correlation of IgA levels and COVID-19 severity (preprint publication).^15^ If it is confirmed that asymptomatic subjects have lower levels of antibodies^30^, this could impact detection of seroconversion in this specific group. We cannot discard that some participants may be either very low or non-responders, as several reports have found COVID-19 patients with low or no responses for IgM, IgG or neutralizing antibodies.^31,32^

By increasing the number of viral antigens in our assay we may allow to increase its sensitivity, as responses to different antigens may present different kinetics and vary between individuals.^33^ Nonetheless, we included determinations to three isotypes to capture a variety of responses between individuals and their relation to time from onset of symptoms. Also, their maintenance and role in protection are probably different. IgM is the first antibody being produced by B cells upon antigenic encounter, IgA is key for mucosal immunity, and IgG is considered to be the most important for memory responses, but their respective kinetics will only be well characterized over a longitudinal follow up study. We found few participants with IgA only, IgM only, or IgG only, and no evidence that specific antibody profiles are associated with the onset of symptoms or current positive rRT-PCR. Therefore, our data do not support that antibody responses could contribute to diagnosis of acute infection (IgM detection +/− increasing IgG) versus past infection (negative or low IgM and persisting IgG) as previously suggested.^34^ However, our analysis of antibody levels and seroprevalence by days since onset of symptoms suggests that IgA responses can be detected and peak earlier than IgM and IgG, consistent with previous reports.^35^

This study has several limitations. First, we collected data over a 12-day period, which, in the context of a rapidly growing epidemic, hinders its association to a specific date, with the prevalence having to be interpreted as the average prevalence over those 12 days. Second, we only collected nasopharyngeal samples (instead of oropharyngeal and nasopharyngeal) from study participants for the molecular detection of SARS-CoV-2 RNA. Although this could reduce rRT-PCR sensitivity, there is evidence showing that nasopharyngeal samples have a higher positivity rate than oropharyngeal samples.^36^ Third, the invalid rRT-PCR results obtained from 43 (7.4%) of the nasopharyngeal swabs analyzed might have led to an under-estimation of overall prevalence of exposure to the virus. Fourth, seroprevalence was defined as positivity of any of the antibody isotypes (IgM, IgG, IgA), which maximized sensitivity rather than specificity. However, our Luminex assay validation showed excellent specificity for the three isotypes, thus, our potential overestimation of the true prevalence is likely to be minimized. Finally, our participation rate (74%) could have introduced selection bias in our sample. It could be that many of those refusing to participate might have had a characteristic associated to an increased risk of infection (being very busy at COVID-19 units, for example). Thus, the impact is potentially minimal, given the lack of association of this and most studied variables with our primary endpoint.

In conclusion, the seroprevalence of antibodies to SARS-CoV-2 was lower than expected. Most participants with a confirmed COVID-19 diagnosis elicited antibody responses (IgA, IgG and/or IgM), with IgA demonstrating the highest sensitivity in the initial days after symptoms onset. Given the current lack of evidence on the correlation of SARS-CoV-2 antibody levels and protection against reinfection, no recommendations should be derived for seropositive HCW at an individual level. This study also shows that around 46% of undiagnosed infections occur in HCW who report having experienced COVID-19 compatible symptoms. Thus, enforcement of rRT-PCR screening programs for all HCW, regardless of the presence of symptoms, is highly recommended in healthcare settings to reduce the risk of hospital-acquired SARS-CoV-2 infections.

## Data Availability

Individual patient data is not available yet.

## Acknowledgements

We would like to thank all the health care workers at Hospital Clínic of Barcelona for their incredible work, endless dedication to patients and resilience. They have given their hearts and health (literally) for people’s health. Special thanks to those HCW who participated in this study, for contributing to expanding our knowledge about COVID-19 immunity and epidemiology. Special thanks to ISGlobal colleagues (Antoni Plasencia, Denise Naniche, Gonzalo Vicente, Matiana González, María Tusell, Cristina Castellana, fellow researchers and administrative department), scientists from Centre for Genomic Regulation (CRG): Juan Valcárcel, Elías Campo from IDIBAPS, and nurses from Occupational Health and Preventive Medicine departments at HCB who have contributed in several ways to make this study happen in the context of such high-pressure clinical care conditions. We are grateful to F. Krammer for donation of RBD protein, to Azucena Bardají for providing the nasopharyngeal swabs, Aida González (Beta implants) for their fantastic face shields, Maria Jesús Mustieles for helping our team of nurses, to Leonie Mayer for assistance with literature review, to Gemma Ruiz-Olalla for statistical advice, to Javier Moreno for assistance with sample processing, to Jordi Vila and Mireia Navarro, for support at the Microbiology Department at HCB.

## Supplementary Material

**Table S1.** Serology results in samples from participants previously diagnosed of COVID-19 by rRT-PCR and more than 10 days since onset of symptoms and pre-pandemic negative controls

| Combination | rRT-PCR+/<br>Sero-<sup>a</sup> | Pre-pandemic/<br>Sero+ | Pre-pandemic/<br>Sero- | rRT-PCR+/<br>Sero+ | Total | Sensitivity (%) | Specificity (%) | PPV (%) | NPV (%) |
| --- | --- | --- | --- | --- | --- | --- | --- | --- | --- |
| **IgG RBD** | 1 | 0 | 47 | 31 | 79 | 97 | 100 | 100 | 98 |
| **IgG RBD &/or IgM RBD** | 1 | 0 | 47 | 31 | 79 | 97 | 100 | 100 | 98 |
| **IgA RBD** | 1 | 1 | 46 | 31 | 79 | 97 | 98 | 97 | 98 |
| **IgA RBD &/or IgG RBD** | 1 | 1 | 46 | 31 | 79 | 97 | 98 | 97 | 98 |
| **IgA RBD &/or IgG RBD &/or IgM RBD** | 1 | 1 | 46 | 31 | 79 | 97 | 98 | 97 | 98 |
| **IgA RBD &/or IgM RBD** | 1 | 1 | 46 | 31 | 79 | 97 | 98 | 97 | 98 |
| **IgM RBD** | 8 | 0 | 47 | 24 | 79 | 75 | 100 | 100 | 85 |

<sup>a</sup> The onset of symptoms of the seronegative participant with positive past rRT-PCR was 19 days before serology testing.
N= 32 participants with positive past rRT-PCR results and 47 pre-pandemic negative controls. PPV, positive predictive value; NPV, negative predictive value.

**Table S2.** Serology results in samples from participants previously diagnosed of COVID-19 by rRT-PCR and in samples from prepandemic negative controls

| **Combination** | **rRT-PCR+/**<br>**Sero–** | **Pre-pandemic/**<br>**Sero+** | **Pre-pandemic/**<br>**Sero–** | **rRT-PCR+/**<br>**Sero+** | **Total** | **Sensitivity (%)** | **Specificity (%)** | **PPV (%)** | **NPV (%)** |
| --- | --- | --- | --- | --- | --- | --- | --- | --- | --- |
| IgA RBD | 6 | 1 | 46 | 33 | 86 | 85 | 98 | 97 | 88 |
| IgA RBD &/or IgG RBD | 6 | 1 | 46 | 33 | 86 | 85 | 98 | 97 | 88 |
| IgA RBD &/or IgG RBD &/or IgM RBD | 6 | 1 | 46 | 33 | 86 | 85 | 98 | 97 | 88 |
| IgA RBD &/or IgM RBD | 6 | 1 | 46 | 33 | 86 | 85 | 98 | 97 | 88 |
| IgG RBD | 7 | 0 | 47 | 32 | 86 | 82 | 100 | 100 | 87 |
| IgG RBD &/or IgM RBD | 7 | 0 | 47 | 32 | 86 | 82 | 100 | 100 | 87 |
| IgM RBD | 14 | 0 | 47 | 25 | 86 | 64 | 100 | 100 | 77 |

N= 39 participants with positive past rRT-PCR samples and 47 pre-pandemic negative controls. PPV, positive predictive value; NPV, negative predictive value.

**Table S3.** Seroprevalence of different antibodies (and combination of) stratified by days since onset of symptoms.

| Combination | Days since onset of symptoms | Sero- | Sero+ | Total | Sero+ (%) |
| --- | --- | --- | --- | --- | --- |
| IgA RBD | 1-9 days | 31 | 2 | 33 | 6.06 |
| IgA RBD | 10-19 days | 47 | 20 | 67 | 29.85 |
| IgA RBD | 20-29 days | 47 | 16 | 63 | 25.4 |
| IgA RBD | 30-39 days | 31 | 2 | 33 | 6.06 |
| IgA RBD | 40-49 days | 7 | 1 | 8 | 12.5 |
| IgA RBD | 50-59 days | 5 | 0 | 5 | 0 |
| IgA RBD | 60-69 days | 1 | 0 | 1 | 0 |
| IgA RBD | No symptoms | 362 | 6 | 368 | 1.63 |
| IgA RBD &/or IgG RBD | 1-9 days | 31 | 2 | 33 | 6.06 |
| IgA RBD &/or IgG RBD | 10-19 days | 46 | 21 | 67 | 31.34 |
| IgA RBD &/or IgG RBD | 20-29 days | 47 | 16 | 63 | 25.4 |
| IgA RBD &/or IgG RBD | 30-39 days | 31 | 2 | 33 | 6.06 |
| IgA RBD &/or IgG RBD | 40-49 days | 7 | 1 | 8 | 12.5 |
| IgA RBD &/or IgG RBD | 50-59 days | 5 | 0 | 5 | 0 |
| IgA RBD &/or IgG RBD | 60-69 days | 1 | 0 | 1 | 0 |
| IgA RBD &/or IgG RBD | No symptoms | 360 | 8 | 368 | 2.17 |
| IgA RBD &/or IgG RBD &/or IgM RBD | 1-9 days | 31 | 2 | 33 | 6.06 |
| IgA RBD &/or IgG RBD &/or IgM RBD | 10-19 days | 45 | 22 | 67 | 32.84 |
| IgA RBD &/or IgG RBD &/or IgM RBD | 20-29 days | 47 | 16 | 63 | 25.4 |
| IgA RBD &/or IgG RBD &/or IgM RBD | 30-39 days | 31 | 2 | 33 | 6.06 |
| IgA RBD &/or IgG RBD &/or IgM RBD | 40-49 days | 7 | 1 | 8 | 12.5 |
| IgA RBD &/or IgG RBD &/or IgM RBD | 50-59 days | 5 | 0 | 5 | 0 |
| IgA RBD &/or IgG RBD &/or IgM RBD | 60-69 days | 1 | 0 | 1 | 0 |
| IgA RBD &/or IgG RBD &/or IgM RBD | No symptoms | 357 | 11 | 368 | 2.99 |
| IgA RBD &/or IgM RBD | 1-9 days | 31 | 2 | 33 | 6.06 |
| IgA RBD &/or IgM RBD | 10-19 days | 46 | 21 | 67 | 31.34 |
| IgA RBD &/or IgM RBD | 20-29 days | 47 | 16 | 63 | 25.4 |
| IgA RBD &/or IgM RBD | 30-39 days | 31 | 2 | 33 | 6.06 |
| IgA RBD &/or IgM RBD | 40-49 days | 7 | 1 | 8 | 12.5 |
| IgA RBD &/or IgM RBD | 50-59 days | 5 | 0 | 5 | 0 |
| IgA RBD &/or IgM RBD | 60-69 days | 1 | 0 | 1 | 0 |
| IgA RBD &/or IgM RBD | No symptoms | 358 | 10 | 368 | 2.72 |
| IgG RBD | 1-9 days | 32 | 1 | 33 | 3.03 |
| IgG RBD | 10-19 days | 47 | 20 | 67 | 29.85 |
| IgG RBD | 20-29 days | 47 | 16 | 63 | 25.4 |
| IgG RBD | 30-39 days | 31 | 2 | 33 | 6.06 |
| IgG RBD | 40-49 days | 7 | 1 | 8 | 12.5 |
| IgG RBD | 50-59 days | 5 | 0 | 5 | 0 |
| IgG RBD | 60-69 days | 1 | 0 | 1 | 0 |
| IgG RBD | No symptoms | 364 | 4 | 368 | 1.09 |
| IgG RBD &/or IgM RBD | 1-9 days | 32 | 1 | 33 | 3.03 |
| IgG RBD &/or IgM RBD | 10-19 days | 45 | 22 | 67 | 32.84 |
| IgG RBD &/or IgM RBD | 20-29 days | 47 | 16 | 63 | 25.4 |
| IgG RBD &/or IgM RBD | 30-39 days | 31 | 2 | 33 | 6.06 |
| IgG RBD &/or IgM RBD | 40-49 days | 7 | 1 | 8 | 12.5 |
| IgG RBD &/or IgM RBD | 50-59 days | 5 | 0 | 5 | 0 |
| IgG RBD &/or IgM RBD | 60-69 days | 1 | 0 | 1 | 0 |
| IgG RBD &/or IgM RBD | No symptoms | 361 | 7 | 368 | 1.9 |
| IgM RBD | 1-9 days | 32 | 1 | 33 | 3.03 |
| IgM RBD | 10-19 days | 51 | 16 | 67 | 23.88 |
| IgM RBD | 20-29 days | 52 | 11 | 63 | 17.46 |
| IgM RBD | 30-39 days | 31 | 2 | 33 | 6.06 |
| IgM RBD | 40-49 days | 7 | 1 | 8 | 12.5 |
| IgM RBD | 50-59 days | 5 | 0 | 5 | 0 |
| IgM RBD | 60-69 days | 1 | 0 | 1 | 0 |
| IgM RBD | No symptoms | 363 | 5 | 368 | 1.36 |

**Figure S1.**
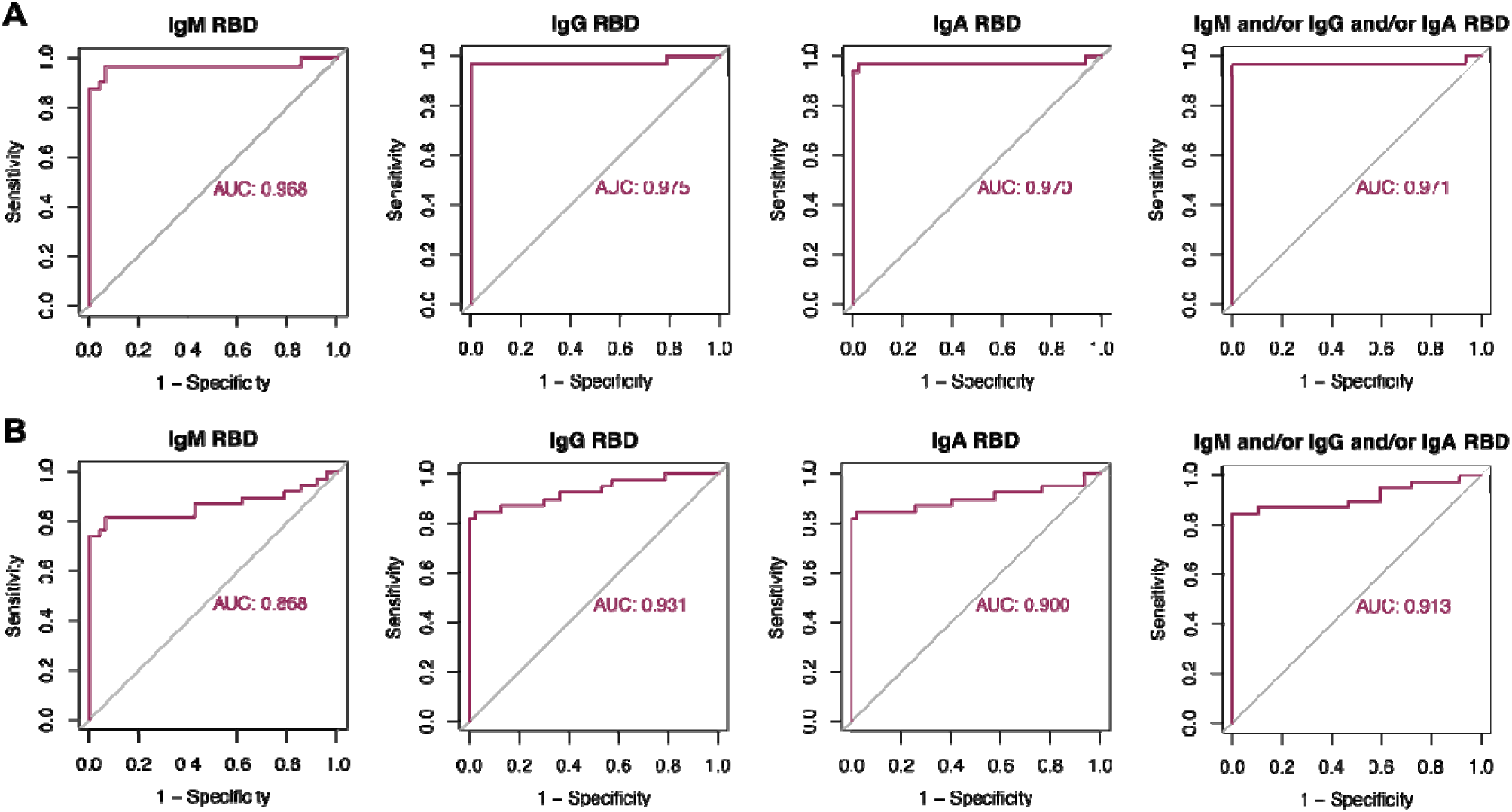
Antibody Luminex assay performance. Receiver operating characteristic (ROC) curve and area under the ROC curve (AUC) for each antibody isotype and the combination of the three isotypes using samples from participants with positive past rRT-PCR and more than 10 days since onset of symptoms (A), and samples from all participants with positive past rRT-PCR regardless of time since onset of symptoms (B). Samples from 32 (A) and 39 (B) study participants and 47 pre-pandemic negative controls were used.

**Figure S2.**
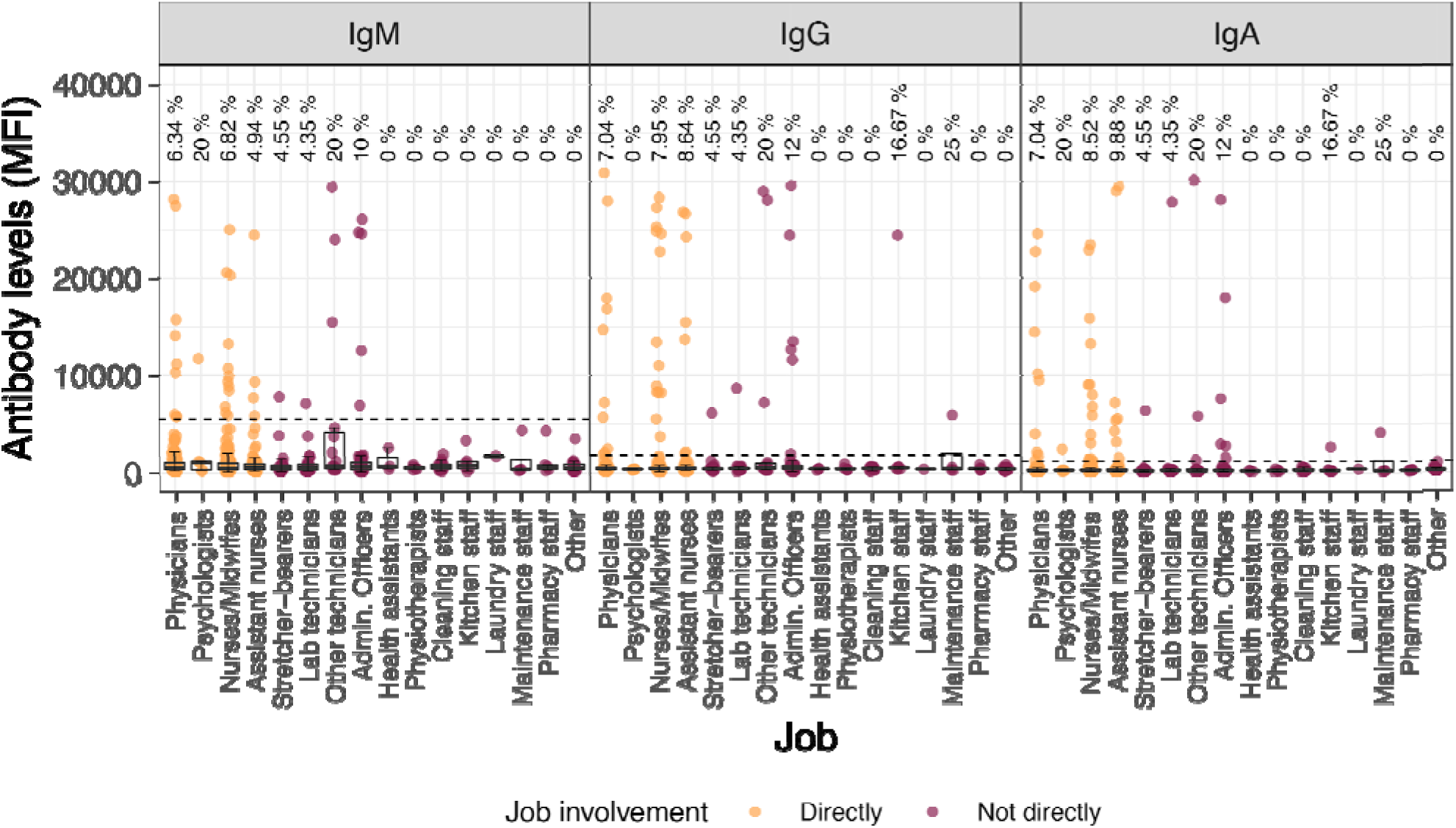
Antibody levels and seroprevalence by professional category. Levels (median fluorescence intensity, MFI) of IgM, IgG and IgA against Receptor Binding Domain of the SARS-CoV-2 Spike glycoprotein by professional role. The dashed line marks the seropositivity threshold. Orange and burgundy dots show subjects working directly with COVID-19 patients or not working directly or not directly with patients, respectively.

